# MultiSurv: Long-term cancer survival prediction using multimodal deep learning

**DOI:** 10.1101/2020.08.06.20169698

**Authors:** Luís A. Vale-Silva, Karl Rohr

## Abstract

The age of precision medicine demands powerful computational techniques to handle high-dimensional patient data. We present MultiSurv, a multimodal deep learning method for long-term pan-cancer survival prediction. MultiSurv is composed of three main modules. A feature representation module includes a dedicated submodel for each input data modality. A data fusion layer aggregates the multimodal representations. Finally, a prediction submodel yields conditional survival probabilities for a predefined set of follow-up time intervals. We trained MultiSurv on clinical, imaging, and four different high-dimensional omics data modalities from patients diagnosed with one of 33 different cancer types. We evaluated unimodal input configurations against several previous methods and different multimodal data combinations. MultiSurv achieved the best results according to different time-dependent metrics and delivered highly accurate long-term patient survival curves. The best performance was obtained when combining clinical information with either gene expression or DNA methylation data, depending on the evaluation metric. Additionally, MultiSurv can handle missing data, including missing values and complete data modalitites. Interestingly, for unimodal data we found that simpler modeling approaches, including the classical Cox proportional hazards method, can achieve results rivaling those of more complex methods for certain data modalities. We also show how the learned feature representations of MultiSurv can be used to visualize relationships between cancer types and individual patients, after embedding into a low-dimensional space.

## Introduction

Millions of cancer deaths are recorded worldwide each year. As incidence and mortality continue to increase, cancer is projected to become the leading cause of death in every country in the world in the 21st century [1]. The prediction of time-to-event outcomes, such as disease recurrence and death, underlies many decisions in the clinical management of cancer patients. Survival analysis, in particular, holds great value for patients, clinicians, researchers, and policy makers [2]. The most basic prognostic technique relies on population-level estimates for the specific cancer site and stage. However, this approach fails to take into account the differences between individual patients, even such fundamental ones as age at diagnosis. To overcome this limitation, a number of patient-specific prognostic methods have been introduced in clinical practice, based on combinations of clinical information and laboratory measurements of validated biomarkers [3]. However, survival predictions still rely often on the clinician’s subjective interpretation and intuition [4], limiting prediction accuracy and reproducibility [5].

In general terms, survival analysis is the study of the time period it takes for an event to occur. In the context of cancer survival, this corresponds to the time between diagnosis and death from the disease. Ideally, survival studies would observe every patient until the target event is recorded. In practice, however, patients are often lost to clinical follow up earlier. In some cases, the event may not even occur at all, if the patient dies from a different cause. Whenever the event of interest is not observed, the last contact time point is referred to as censoring time. Censored observations still contain useful information for modeling, however, since the censoring time provides a lower bound on the patient’s survival time. The classical statistical approach to model survival data with censored observations is the semi-parametric Cox proportional hazards (CPH) model [6]. This method is used very widely and with great success [7], but it has two important constraints that may be unrealistic in practice. On the one hand, CPH is based on a linear model, making it unable to capture non-linear relationships between the input data and the risk of death. On the other hand, it assumes that the effect of the patient’s features is constant over time, constraining the method to yield proportional patient predictions at all follow up time points.

A successful approach to overcome CPH’s linearity constraint has been to use Deep Learning (DL) models. Deep Learning, a subfield of Machine Learning, uses artificial neural networks to discover representations from the raw input data automatically, bypassing the need for manual feature engineering of traditional machine learning [8]. Deep neural networks have the ability to model highly complex non-linear relationships and have already demonstrated breakthrough success in healthcare [9] and beyond [8]. The first approach using DL models within the CPH framework consisted of a simple feed-forward neural network for unimodal data [10]. The advent of big data collection in healthcare for precision medicine resulted in a wealth of new data modalities being increasingly available in routine clinical practice. Integration of such big data demands powerful modeling approaches, reinforcing the call for DL-based methods [9, 11]. Accordingly, a number of recent studies leveraged modern DL techniques to increase the capacity of DL-based CPH [12]. Notable examples include methods designed to use clinical and gene expression data, namely DeepSurv [13] and Cox-nnet [14]. Other methods focused on imaging data, such as CXR-risk, which uses chest radiographs [15], and WSISA, which employs histopathology slides [16].

The current availability of high-dimensional *multimodal* data, including clinical, imaging, and high-throughput molecular data, calls for their integration within the framework of deep multimodal representation learning [17, 18]. Several recent studies have extended DL-based Cox survival methods to integrate different data modalities and allow more accurate predictions. One notable example is SurvivalNet, which uses different high-throughput molecular data modalities from different cancer types [19]. The GSCNN system combines digital pathology images with two validated genomic biomarkers from glioma patients [20]. SALMON addresses breast cancer using a combination of gene and microRNA expression with a handful of clinical parameters and validated biomarkers [21]. More recently, this approach was extended to use four different data modalities, including clinical information, digital pathology images, and two different genomics data modalities (gene and micro RNA expression), integrated in a multimodal DL model for pan-cancer prognosis prediction across 20 cancer entities [22]. While these methods overcome CPH’s linearity constraint using non-linear DL models, they still yield proportional hazards.

Recently, methods have been proposed to overcome both the linearity and the proportionality constraints of the CPH model. One method, named Cox-Time, handles time as an additional input feature to model its interactions with the regular input features [23]. Other methods employ a fully-parametric approach, relying on discretization schemes of the measured time and outputting predictions for a set of predetermined time intervals. One method uses multi-task logistic regression [24], while a related method, named Dynamic-DeepHit, parameterizes the probability mass function of the survival distribution and adds a ranking component to the loss [25]. Another approach consists in parameterizing a discrete conditional hazard rate at each time interval. This method was originally developed decades ago [26], but a recent version, called Nnet-survival, has adapted it to leverage modern DL techniques [27]. Very recently, a completely different approach has been proposed, consisting in the transformation of survival times into jackknife pseudo conditional survival probabilities [28]. This casts survival prediction as a standard regression problem. These latter approaches [23-28] address both main constraints of CPH (linearity and hazard proportionality) but do not exploit the available multimodal data types.

In this work, we introduce MultiSurv, an end-to-end <u>multi</u>modal DL-based and discrete-time <u>surv</u>ival prediction method for pan-cancer patient prognosis estimation. This method extends our MultiSurv prototype recently presented at a conference [29]. While both MultiSurv versions overcome the linearity constraint by using non-linear DL-based models, the new MultiSurv method introduced here uses a different data fusion layer and, additionally, overcomes the proportionality constraint using the Nnet-survival approach [27]. Thus, the proposed MultiSurv method is the first non-linear and non-proportional method to handle multimodal data. MultiSurv delivers highly accurate long-term survival predictions for patients diagnosed with a wide variety of cancer types. We compared MultiSurv with previous methods, including classical CPH and DL-based nonlinear and non-proportional methods. With unimodal data, the methods yielded partially competitive results, but MultiSurv achieved the best performance. Integrating multimodal data further increased the performance of MultiSurv.

## Results

### DL-based multimodal method for survival prediction: MultiSurv

MultiSurv has a modular architecture, with dedicated input data modality submodels, a data fusion layer, and a final survival prediction fully-connected neural network submodel. MultiSurv determines a conditional survival probability for each time interval in a predefined set of follow-up time intervals. A schematic overview of the model architecture is presented in Fig. 1. MultiSurv uses a set of six data modalities with potential complementary predictive value in cancer survival. These include clinical and demographic information, multi-omics data from four different modalities, and tissue biopsy imaging data. The multi-omics modalities consist of genomics (copy number variation), transcriptomics (gene and microRNA expression) and epigenomics (DNA methylation) data, potentially containing established or novel biomarkers [31]. The imaging data, on the other hand, contain tissue architecture information [32] that is lost in bulk-analysis omics data. Since DL models can handle raw data and automate feature engineering, we used only relatively simple feature selection techniques to reduce computational cost. In order to make full use of the available dataset, MultiSurv can also cope with patients with missing data. Missing values within single data modalities are handled using standard techniques: median substitution for continuous features, and introduction of an additional category for categorical features. Completely missing data modalities are also handled seamlessly using a dropout mechanism, with MultiSurv relying on the available modalities (see Methods section for further details).

**Fig. 1.**
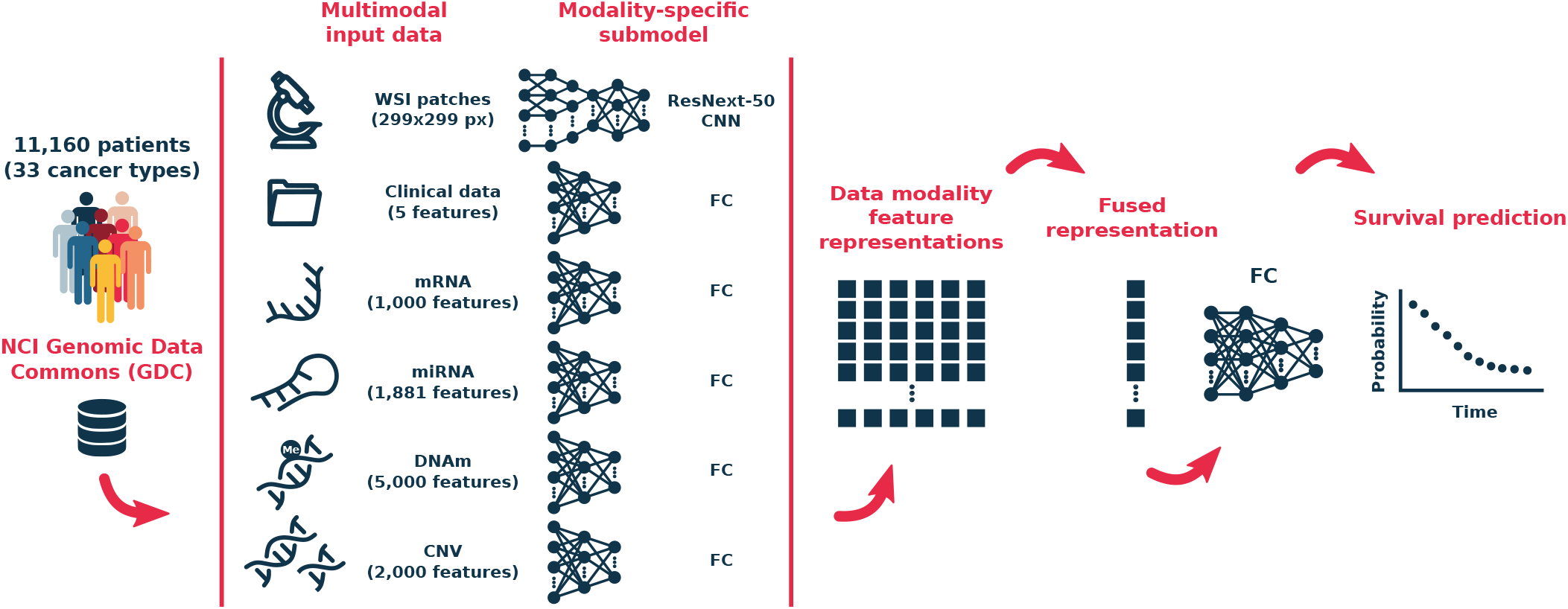
The MultiSurv model architecture. Input data are all from the NCI Genomic Data Commons database, including up to six different data modalities. Each data modality is handled by a dedicated DL submodel, trained to generate modality-specific feature representations. A data fusion layer combines the generated feature representation vectors into a single fused representation. A final neural network takes the fused feature representation and outputs a conditional survival probability for each of a set of pre-defined follow-up time intervals. Taking the cumulative product of the set of conditional survival probabilities produces the predicted survival curve. CNN: convolutional neural network; FC: fully-connected neural network.

In MultiSurv, each input data modality is handled by a dedicated submodel, which automatically determines an appropriate representation of the input features. For the clinical and omics data modality submodels, we used a fully-connected neural network architecture with up to five hidden layers, a rectified linear unit (ReLU) non-linear activation function, dropout regularization, and batch normalization. For the imaging modality submodel we used a ResNeXt-50 convolutional neural network [33] pretrained on the ImageNet natural image dataset [34] and fine-tuned within end-to-end training of MultiSurv. All data modality submodels output a feature representation vector of length 512. The data fusion layer integrates the multimodal feature representations by taking the element-wise maxima across the set of representation vectors, reducing them to a single fusion vector of the same length. This fusion vector is the input to a fully-connected neural network with 30 output units, one for each time interval of a set of predefined time intervals. MultiSurv yields predictions for time intervals of one year (spanning a combined total follow-up time of 30 years), each predicting the respective conditional survival: the probability of surviving that time interval given that the subject has survived at least to the beginning of the interval. The number and time length of the discrete output intervals is very flexible, since different choices achieve similar accuracy Fig. S1). The configuration of the output layer, the different modular components, and the tunable parameters was determined empirically. For more details on MultiSurv’s architecture we refer the reader to the Methods section.

### MultiSurv achieves high prognostic accuracy

We measured the accuracy using two different metrics: the time-dependent concordance index [35], referred to as C^td^, and the integrated Brier score [36], abbreviated as IBS. The C^td^ is an extension of Harrell’s concordance index (also known as C-index), a nonparametric statistic that quantifies the ability of the predictive model to discriminate among subjects with different event times [37]. Just like the C-index, the C^td^ is a measure of the model’s discrimination power. A C^td^ of 1 indicates perfect concordance between predicted risk and actual survival, while a value of 0.5 means random concordance. The IBS, on the other hand, is based on the average squared distances between observed survival status and predicted survival probabilities at all available follow up times. It extends the Brier score, which applies to a single time point. The IBS measures both the discrimination power and the calibration of the model’s predictions. The lower the IBS, the better the model performance, with the best value at zero.

We started by evaluating MultiSurv’s performance with unimodal data, in order to validate the modality feature extractors, as well as to compare MultiSurv to existing methods that cannot handle multimodal data. We considered the classic CPH model [6], random survival forests [30], and two non-linear DL-based methods: the proportional hazard method DeepSurv [13], and the non-proportional Deep-Hit [25]. As can be seen in Table 1, MultiSurv achieved the best results for almost all data modalities for both metrics. Exceptions were gene expression data (mRNA), for which MultiSurv obtained the second best IBS score (after CPH), and DNA methylation data, for which it obtained a C^td^ score just below the second best, while still displaying the best IBS score. Among the six data modalities, the highest performance was obtained for clinical data, while imaging data (WSI) yields the lowest performance

**Table 1.** Method performance with unimodal data inputs. Time-dependent concordance index (C^td^) and integrated Brier score (IBS) with 95% bootstrap confidence interval (CI; numbers in parentheses). The best and second best results for each metric for each data modality are boldfaced and underlined, respectively.

| Data | Metric | Method |  |  |  |  |
| --- | --- | --- | --- | --- | --- | --- |
|  |  | CPH [6] | RSF [30] | DeepSurv [13] | DeepHit [25] | MultiSurv |
| Clinical | $C^{td}$ | <u>0.796</u> (0.779-0.813) | 0.770 (0.751-0.789) | 0.792 (0.773-0.810) | **0.809** (0.792-0.826) | **0.809** (0.793-0.825) |
|  | IBS | **0.143** (0.135-0.154) | 0.184 (0.179-0.191) | **0.143** (0.134-0.154) | <u>0.173</u> (0.165-0.184) | **0.143** (0.134-0.155) |
| mRNA | $C^{td}$ | 0.733 (0.712-0.755) | 0.719 (0.695-0.741) | 0.746 (0.722-0.768) | <u>0.752</u> (0.728-0.774) | **0.758** (0.735-0.780) |
|  | IBS | **0.177** (0.165-0.190) | 0.191 (0.181-0.200) | 0.180 (0.159-0.198) | 0.191 (0.180-0.198) | <u>0.178</u> (0.157-0.194) |
| DNAm | $C^{td}$ | <u>0.739</u> (0.719-0.760) | 0.729 (0.709-0.752) | **0.759** (0.739-0.780) | 0.737 (0.715-0.758) | 0.736 (0.714-0.759) |
|  | IBS | 0.179 (0.165-0.192) | 0.186 (0.176-0.192) | <u>0.177</u> (0.156-0.203) | 0.194 (0.179-0.208) | **0.175** (0.156-0.189) |
| miRNA | $C^{td}$ | 0.676 (0.651-0.700) | 0.664 (0.639-0.689) | 0.685 (0.661-0.711) | <u>0.700</u> (0.674-0.725) | **0.702** (0.677-0.728) |
|  | IBS | <u>0.186</u> (0.171-0.202) | 0.193 (0.183-0.201) | 0.194 (0.170-0.219) | <u>0.186</u> (0.177-0.197) | **0.179** (0.160-0.199) |
| CNV | $C^{td}$ | 0.570 (0.543-0.599) | <u>0.604</u> (0.579-0.630) | 0.596 (0.571-0.621) | 0.575 (0.549-0.599) | **0.617** (0.591-0.643) |
|  | IBS | <u>0.214</u> (0.207-0.224) | 0.217 (0.208-0.225) | 0.229 (0.215-0.247) | 0.217 (0.212-0.224) | **0.210** (0.200-0.221) |
| WSI | $C^{td}$ | - | - | - | - | **0.569** (0.543-0.597) |
|  | IBS | - | - | - | - | **0.220** (0.206-0.231) |

CPH: Cox Proportional Hazards; RSF: Random Survival Forest; Clinical: tabular clinical data; mRNA: gene expression; DNAm: DNA methylation; miRNA: microRNA expression; CNV: gene copy number variation; WSI: whole-slide images.

We also evaluated MultiSurv using multimodal input data, with different numbers and combinations of the six data modalities. Table 2 lists the combinations yielding the best performance. Overall, judging by the results of different data modality combinations, the contribution of each data modality to the multimodal models corresponds to the results in the unimodal configuration. Accordingly, including clinical data turned out to be necessary to achieve best results in the multimodal case. The highest performance was obtained with bi-modal inputs combining clinical data with gene expression (mRNA; highest C^td^ value) or DNA methylation (DNAm; highest IBS). Combining more than two modalitites resulted in slightly lower performance. To facilitate end-to-end training of the multimodal configurations, we investigated leveraging the available trained unimodal models. We used the model weights from pretrained unimodal clinical and mRNA models to initialize the respective submodel weights of the bimodal clinical and mRNA MultiSurv configuration. While, as expected, this approach allowed faster convergence, it did not yield performance improvements. In addition, we tested a data dropout scheme, consisting of dropping a random data modality from each patient with a predefined probability during training. However, this did not yield improvements either. Multimodal data dropout rates of up to 0.25 (25% probability of dropping one data modality for each patient during model training) had no impact in MultiSurv’s results, while higher rates led to decreases in performance.

**Table 2.** Model performance using a selection of combinations of the six input data modalities. Individual data modalities included in each evaluated model are marked with •. The best and second best results for each metric are boldfaced and underlined, respectively.

| Included data modalities | | | | | | $C^{td}$<br>(95% CI) | IBS<br>(95% CI) |
| --- | --- | --- | --- | --- | --- | --- | --- |
| Clinical | mRNA | DNAm | miRNA | CNV | WSI |  |  |
| • | • |  |  |  |  | **0.822** | <u>0.138</u> |
|  |  |  |  |  |  | (0.805-0.837) | (0.126-0.150) |
| • |  | • |  |  |  | 0.808 | **0.134** |
|  |  |  |  |  |  | (0.791-0.826) | (0.125-0.148) |
| • |  |  | • |  |  | 0.792 | 0.147 |
|  |  |  |  |  |  | (0.775-0.810) | (0.136-0.161) |
| • |  |  |  | • |  | 0.795 | 0.140 |
|  |  |  |  |  |  | (0.778-0.812) | (0.131-0.152) |
| • |  |  |  |  | • | 0.801 | 0.148 |
|  |  |  |  |  |  | (0.783-0.817) | (0.140-0.158) |
| • | • | • |  |  |  | <u>0.810</u> | 0.146 |
|  |  |  |  |  |  | (0.793-0.829) | (0.135-0.158) |
| • | • | • | • |  |  | 0.798 | 0.153 |
|  |  |  |  |  |  | (0.781-0.815) | (0.139-0.168) |
| • | • | • | • | • |  | 0.802 | 0.149 |
|  |  |  |  |  |  | (0.748-0.820) | (0.136-0.162) |
| • | • | • | • | • | • | 0.787 | 0.152 |
|  |  |  |  |  |  | (0.769-0.806) | (0.140-0.166) |

Clinical: tabular clinical data; mRNA: gene expression; DNAm: DNA methylation; miRNA: microRNA expression; CNV: gene copy number variation; WSI: whole-slide images.

Concerning individual cancer types, we found a relatively large variability of the results. We analysed cancer types with at least 20 patients in the test dataset (Fig. S2) and found C^td^ values approaching the optimal score of 1.0 for thyroid carcinoma (THCA; 0.988), kidney renal papillary cell carcinoma (KIRP; 0.959), and colon adenocarcinoma (COAD; 0.953). For sarcoma (SARC) and lung squamous cell carcinoma (LUSC), on the other hand, relatively low scores were obtained (0.589 and 0.554, respectively). For the IBS metric similar results were found. The best results were obtained for THCA (0.045), KIRP (0.066), and prostate adenocarcinoma (PRAD; 0.079). The worst scores were obtained with SARC (0.265), head and neck squamous cell carcinoma (HNSC; 0.225), and LUSC (0.224).

### MultiSurv predicts long-term survival

MultiSurv’s multiple discrete-time output layer yields time-varying conditional survival predictions. These can be used to generate patient survival curves for the covered time span. We trained MultiSurv with yearly output time points up to 30 years from diagnosis, corresponding to the latest event time recorded in the training dataset. The latest event time in the test dataset is around 20 years, so we display MultiSurv’s predictions up to that time point. We started by visualizing the predicted survival curves for four cancer types with different prognoses: prostate adenocarcinoma (PRAD), with very good prognosis; kidney renal clear cell carcinoma (KIRC) and ovarian serous cystadenocarcinoma (OV), with worse prognosis; and glioblastoma multiforme (GBM), with very poor prognosis. Survival curves predicted by MultiSurv illustrate these differences very well (Fig. 2a). Patient group curves by cancer type can be obtained by averaging all respective patient predictions. These can then be compared with the survival curves constructed using the Kaplan-Meier estimator, a non-parametric statistic used to estimate the survival function and visualize the actual survival trend in the available data. As shown in Fig. 2b, predicted cancer type curves follow the Kaplan-Meier estimates very closely. The same is true of the overall survival curve for the complete test dataset (Fig. 2c). MultiSurv predictions for all 30 other cancer types can be found in Fig. S3. MultiSurv can thus be used to generate accurate long-term predictions for previously unseen patients. In order to test this more formally, we used MultiSurv to identify two different risk groups. We split the patients in the test dataset into two groups according to their survival probabilities predicted by MultiSurv. This yielded risk groups that do indeed have significantly different survival distributions (log-rank *p* value 2.3^-56^ comparing Kaplan-Meier estimates; Fig. 2d), confirming the accuracy of the model’s predictions.

**Fig. 2.**
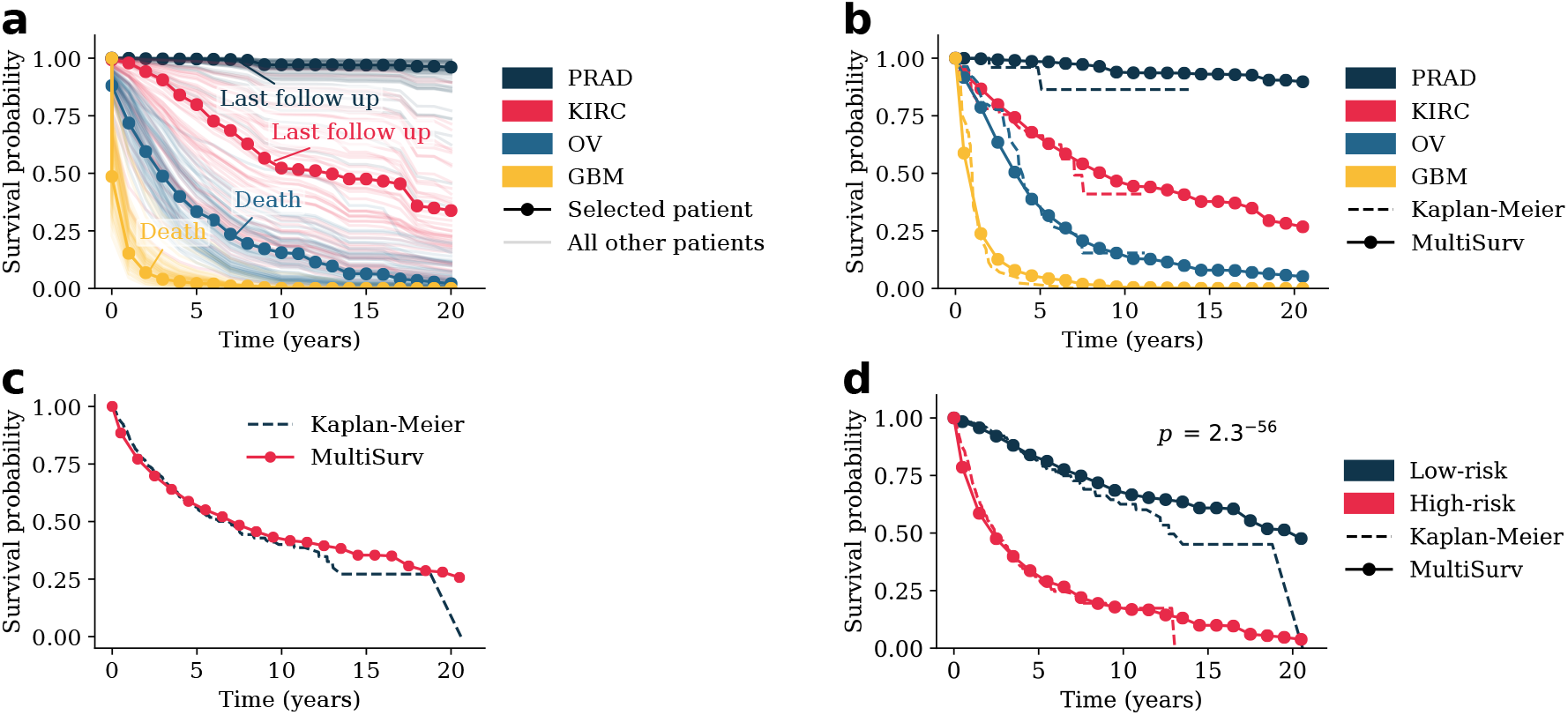
MultiSurv predictions allow construction of accurate survival curves. MultiSurv outputs patient survival predictions for the defined discrete-time follow up intervals. These can then be averaged to obtain group-wide survival predictions. **a)** Survival curves constructed using Multisurv predictions for each patient in the test dataset diagnosed with one of four selected cancer types. One example patient is highlighted for each cancer type and the corresponding last follow up time point is annotated (as “Last follow up” if the patient is censored or “Death” if it corresponds to patient death). Highlighted patient codes are TCGA-HI-7169 for PRAD, TCGA-B0-5691 for KIRC, TCGA-29-1762 for OV, and TCGA-19-1390 for GBM. **b)** Survival curves for the four example cancer types in panel a) compared with Kaplan-Meier estimator outputs. **c)** Survival curves for all patients in the test dataset compared with the Kaplan-Meier estimator output. **d)** MultiSurv predictions allow accurate stratification of patient risk groups. Patients were split into low and high-risk groups according to MultiSurv's first output interval risk prediction using the median value across all patients as the threshold. The two resulting groups have significantly different Kaplan-Meier estimates (log-rank test). The plot shows MultiSurv prediction averages overlayed on the Kaplan-Meier estimators. PRAD: Prostate Adenocarcinoma; KIRC: Kidney Renal Clear Cell Carcinoma; OV: Ovarian Serous Cystadenocarcinoma; GBM: Glioblastoma Multiforme.

### MultiSurv yields non-proportional predictions

The learned effect of the input data on MultiSurv’s survival probability predictions for each output time interval can vary freely. In other words, MultiSurv is not constrained by the proportional hazards assumption of Coxian methods (both the classic linear method and its non-linear developments). This is more realistic, since many input features in the data used in this study seem to violate the proportional hazards assumption. Testing the assumption that the influence of the input features is constant over time, which underlies the hazard proportionality in the CPH family of models, suggests that for many features in the data it does not hold. For example, for six out of a total of 10 features in the clinical data modality (race, prior malignancy, pharmaceutical treatment, radiation treatment, tumor stage, and age at diagnosis) the null hypothesis of non-varying effects is rejected *(p <* 0.05). Similarly, out of the 50 principal components of each omics data modality, 12, 18, 12, and 13 of mRNA, DNAm, miRNA, and CNV data, respectively, all fail the test as well. Additionally, it is easy to find examples of crossed survival curves in the data (Fig. S4). These cannot be reproduced by methods constrained by the proportional hazard assumption, which output patient predictions with the same ranking for all follow up time points.

### MultiSurv learns effective feature representations

In order to gain insights towards the network’s internal representation of the patient data, we investigated the feature representations learned by MultiSurv. We focused on the fused multimodal representation vector in the model with the highest C^td^ value, the configuration with clinical and mRNA input data. We used the t-distributed Stochastic Neighbor Embedding algorithm (t-SNE) [38] to embed the multimodal fused representations generated for each patient in the test dataset from the original 512-dimensional space into a twodimensional space (Fig. 3a). To inform the visualization, we highlighted points for patients diagnosed with each of the previously selected cancer types. The remaining cancer types are visualized in Fig. S5. Patients diagnosed with each of the cancer types appear in specific clusters and occupy different regions of the two-dimensional space, aligning well with the known cancer type prognosis. In the particular instance of the embedding visualized here, moving along the twodimensional space from left to right corresponds roughly to the progression from the good prognosis of PRAD, through the intermediate prognoses of KIRC and OV, to the very poor prognosis of GBM (compare the corresponding survival curves in Fig. 2b). Patients diagnosed with PRAD mostly occupy a tight cluster. Similarly, GBM patients even form their own island, indicating highly specific characteristics of this cancer type. Patients diagnosed with KIRC and OV give rise to more heterogeneous representations (Fig. 3a), hinting at the existence of distinct subpopulations within those cancer types. These learned representations are also useful to identify survival outliers. We picked a few patients whose embedded representations stand out visually from their cancer type clusters and plotted their predicted survival curves. This allowed convenient identification of some of the patients with most extreme prognosis within their respective cancer type (Fig. 3b-e). The distribution of individual patient representations within cancer types also serves to visualize each cancer type’s heterogeneity in the distribution of survival predictions. The cancer types with embedded points closely clustered together, PRAD and GBM, show relatively homogenous patient prognosis (Fig. 3b and d). On the contrary, KIRC and OV appear as less defined clusters in Fig. 3a, with correspondingly larger variation in prognosis (Fig. 3c and e).

## Discussion

We developed the DL-based multimodal survival prediction method MultiSurv, the first non-linear and nonproportional method to handle multimodal data inputs. We investigated the combination of clinical information, digital pathology images, and several high-dimensional genomic data modalities from patients diagnosed with one of 33 different cancer entities, publicly available from the National Cancer Institute’s (NCI) Genomic Data Commons (GDC) database. The network, including the data modality-specific feature representation submodels, the multimodal fusion layer, and the discrete-time survival predictor, was trained end-to-end using stochastic gradient descent. MultiSurv delivers non-proportional outputs and achieves high accuracy in long-term prediction across cancer entities. Overall, MultiSurv achieved the best scores for virtually every tested data modality, with clinical data proving to be the most informative (Table 1). Using the multimodal input configuration allowed further increases in performance for specific data modality combinations (Table 2).

For unimodal data, we found that the compared baseline methods generally showed good performance. Even the classical CPH method achieved partially competitive results. The reason for this may be the fact that the number of available patients in the dataset (around 11,000) is relatively limited for DL methods. We speculate that the increasing availability of patient data in the future will likely lead to a larger performance difference between classical and DL-based methods. Likewise, further benefit of using multimodal inputs is to be expected, which may favor MultiSurv’s superiority even further. With the currently available dataset, however, classical CPH using tabular clinical data can be considered a good choice, particularly when specialized computer hardware required for DL is not available and some drop in performance can be tolerated.

Recent work by Cheerla and Gevaert [22] also tackled pan-cancer survival prediction using a DL-based approach. Their method relies on a DL-based CPH model, as in Deep-Surv [13], extended to handle multimodal data. The authors used the same TCGA database employed here, but included only 20 of the 33 available cancer types. They reported a best pan-cancer C-index of 0.784 (the values for the C-index and C^td^ are the same, since the method outputs proportional hazards). MultiSurv yielded a better performance, with a best C^td^ of 0.822 using all 33 cancer types. To compare the results in [22] more directly with those of MultiSurv, we reduced the dataset to the same 20 cancer types (in our setting 9,197 patients were included out of the total 11,081) and trained MultiSurv on clinical and mRNA data inputs. With this configuration, MultiSurv still achieved a higher C^td^ of 0.801. We also noted interesting differences in the most informative data modalities. Cheerla and Gevaert reported miRNA and mRNA as the most and least informative modalities, respectively. The reason behind the lower value of the clinical data modality in that study is particularly interesting and is probably due to the fact that only four of the available clinical features were used. We found that the additional features used in our work, even if missing in a considerable percentage of the patients, contribute to improved performance. Beyond that, the differences in performance may be explained by differences in model architecture between that model and MultiSurv.

MultiSurv generally shows better performance than other previous multimodal methods trained on TCGA datasets as well, with the exception of the results of SurvivalNet [19] and GSCNN [20] for glioma patients. These studies report evaluations on combined data from glioblastoma multiforme (GBM) and brain lower grade glioma (LGG), with C-index values above 0.8 (higher than MultiSurv’s C^td^ of 0.650 and 0.741, respectively). Like the pan-cancer method described above, these other previous methods yield proportional hazards. In any case, MultiSurv’s predictions are still very well calibrated (particularly for GBM, as evidenced by the very low IBS score of 0.109). The relatively low C^td^ values can be explained by the fact that survival curves for glioma patients are very similar across patients (see predicted curves for GBM patients in Fig. 3d), allowing for a well calibrated model to still yield incorrect rankings of patient survival probabilities (measured by the C-index and C^td^ metrics).

MultiSurv could be further improved in several ways. One main avenue would be to include additional input features and additional data modalities. In MultiSurv, this would require only slight adjustments to the current architecture and the development of additional dedicated submodels, respectively. Additional feature representation submodels can be seamlessly integrated into the overall architecture. Another avenue concerns the digital pathology image submodel. Judging from the value this data provides to pathologists, MultiSurv’s current WSI submodel could be improved. The main challenge is coping with the wide variety of tissue appearances in over 30 cancer types (from a comparably wide variety of physiological systems), as well as the large size of the input images (gigapixel-level digitized slides). More sophisticated patch sampling techniques might improve the result. Finally, even though we tested a wide range of different techniques already, we believe there is potential to improve MultiSurv’s multimodal representation learning approach. It would be useful to explore coordinated representation learning alternatives, for example, as well as early and late fusion frameworks [17, 18].

In conclusion, in this study we developed MultiSurv, a non-linear, non-proportional hazard discrete-time pan-cancer survival prediction system. The best model architecture was determined by investigating a wide variety of data modalities, as well as multimodal data fusion techniques. MultiSurv learns effective internal representations of the raw multimodal input data, which allows accurate long-term survival predictions for patients diagnosed with a wide variety of cancer types. MultiSurv can leverage the multiple high-dimensional data modalities now available in precision medicine-based clinical practice. This way, MultiSurv can be a useful tool in the clinical management of cancer patients, helping clinicians deliver accurate and reproducible prognosis predictions.

**Fig. 3.**
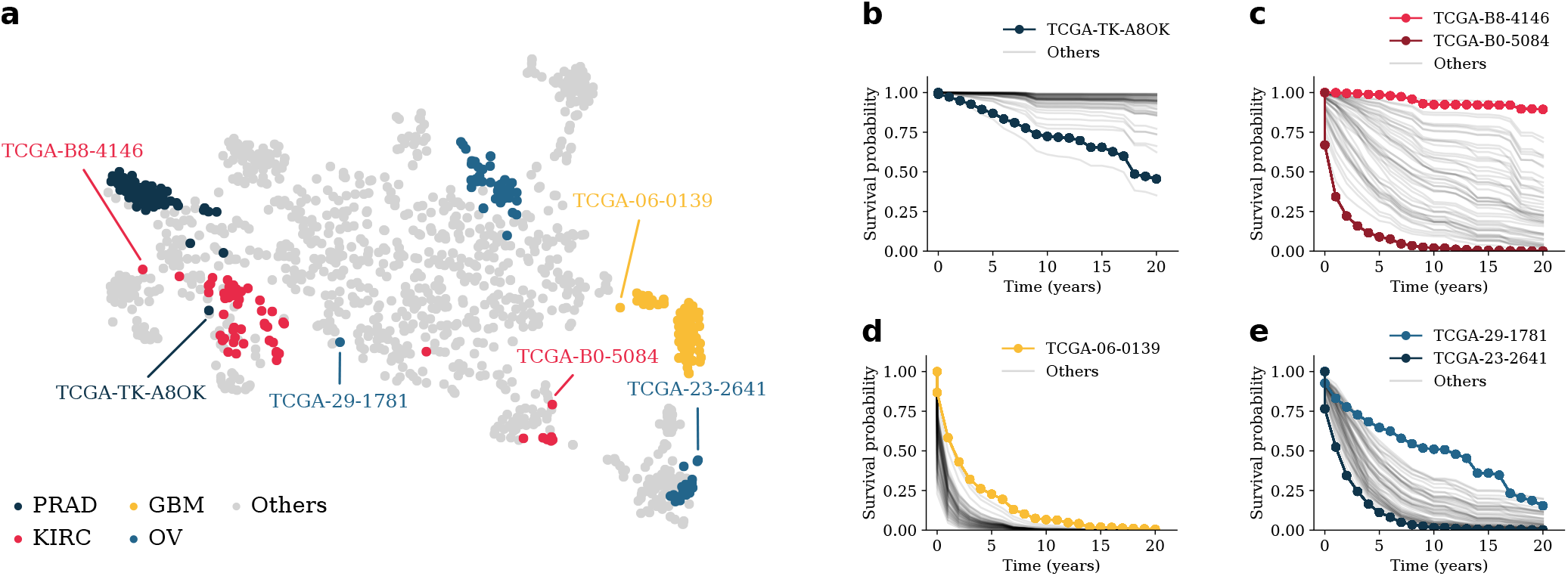
Visualization of feature representations learned by MultiSurv. We collected the internal fused feature representation vector of the MultiSurv model trained on clinical and mRNA data and embedded it into a two-dimensional space using t-SNE. **a)** Embedded feature representations for each patient in the test dataset. Patients diagnosed with each of four selected cancer types are highlighted. Within each of the highlighted cancer types, visually selected outlier patients are annotated. All patient survival curves, highlighting the selected outliers, are displayed for **b)** PRAD, **c)** KIRC, **d)** GBM, and **e)** OV. PRAD: Prostate Adenocarcinoma; KIRC: Kidney Renal Clear Cell Carcinoma; OV: Ovarian Serous Cystadenocarcinoma; GBM: Glioblastoma Multiforme.

## Methods

### Data

All data used in this work are from the National Cancer Institute (NCI) Genomic Data Commons (GDC) information system [39], publicly accessible at the GDC Data Portal (https://portal.gdc.cancer.gov/). We used the data subset produced by The Cancer Genome Atlas (TCGA) program, which includes a rich body of imaging, clinical, and molecular data from 11,315 cases of 33 different cancer types [40]. Patients were followed for a recorded length of time until death or loss to clinical observation. We downloaded the clinical data table for the TCGA project using the TCGAbiolinks package v2.8.4 in the R statistical computing software environment v3.5.1. This table includes patient codes, available clinical features, as well as survival labels (“vital_status”, corresponding to the event indicator, and follow up durations: “days_to_last_follow_up” and “days_to_death”) for a total of 11,167 patients. We dropped a few additional patients with incomplete label information: 16 patients missing vital status information; 11 patients with recorded death but missing “days_to_death”; and 53 patients missing both follow up durations. The final number of patients, percentage of censored observations, and simple descriptive statistics of follow up durations for each cancer type are listed in Table S1.

We used six different data modalities: tabular clinical data (herein simply referred to as "clinical"), gene expression (referred to as "mRNA"), microRNA expression (miRNA), DNA methylation (DNAm), gene copy number variation (CNV) data, and whole-slide images (WSI). We performed some feature selection and data pre-processing for each data modality. Details of the different data modalities, as well as data pre-processing procedures, are provided in Suppl. Note 1. The final number of patients and features in each data modality after preprocessing is listed in Table 3.

**Table 3.** Summary information of the different data modalities after preprocessing.

| Modality | No. patients | No. features |  |
| --- | --- | --- | --- |
|  |  | Continuous | Categorical |
| Clinical | 11,081 | 1 | 9 |
| mRNA | 9,605 | 1,000 | - |
| DNA<sub>m</sub> | 10,257 | 5,000 | - |
| miRNA | 9,616 | 1,881 | - |
| CNV | 10,325 | - | 2,000 |
| WSI | 8,376 | 299 × 299 | - |

MultiSurv can handle missing data, which allows full use of the available dataset. Many patients lack entries for specific features of the clinical data modality. This is handled in the data preprocessing stage as described in Suppl. Note 1 (section "Tabular clinical data"). When training MultiSurv with unimodal data, patients missing the entire respective data modality are excluded. For multimodal data, single missing data modalities are coped with by replacing them by a zero input of the same dimension, to allow integration in the patient batching procedure. This allows the model to learn from existing data of other modalities. MultiSurv also includes a multimodal data dropout option to avoid overfitting, implemented using the same mechanism. Concretely, during model training, one data modality from each patient is chosen at random with a specified probability and replaced by a zero input (provided at least two modalities are available for the patient).

### Validity of proportional hazards assumption

The proportional hazards assumption is a core restriction of the CPH family of models. These models include a non-parametric time-varying baseline hazard that is equal for all study subjects. The baseline hazard is scaled by a factor determined by a parametric term that depends on the subjects’ input features, but which is constant over time. In other words, these models assume that the effect of the input features does not vary over time. The predicted survival probabilities for any two given subjects are thus proportional to each other at all considered time points. We tested the validity of the proportional hazard assumption for the data used in this work using a statistical test for time-varying feature effects. We fit a CPH model on single data modalities from the training data using the lifelines software library v0.23.8 [41]. As for model evaluation, we used all features when modeling clinical data and the 50 principal components when modeling omics data modalities. We then used the "check_assumptions" method to run the statistical test and rejected the null hypothesis (feature effect does not vary with time) for features with a *p* value below 0.05. Additionally, we inspected plots of scaled Schoenfeld residuals [42] for all features for which the null hypothesis is rejected, which are a part of the output of the method and provide a visualization of the time-varying effects.

### Model architecture

MultiSurv is a deep multimodal discrete-time survival prediction system with a modular architecture as shown in Fig. 1. The overall architecture is composed of three core modules: a feature representation module, consisting of dedicated data modality submodels, each outputting a fixed-size hidden data representation; a multimodal data fusion layer, which fuses the data modality submodel outputs into a single representation; and an output submodel that maps the incoming fused feature representation to a set of discrete-time conditional survival probability predictions. We used fully-connected neural networks with two to five hidden layers, the rectified linear unit (ReLU) nonlinear activation function, dropout regularization, and batch normalization for all data modalities except imaging data (WSI). The dedicated WSI submodel consists of a ResNeXt-50 [33] convolutional neural network pre-trained on the ImageNet natural image dataset [34]. For integration in MultiSurv, we replaced ResNeXt-50’s fully-connected output layer by a 512-unit layer, to match the fixed size of MultiSurv’s data modality feature representations. For training, we fixed ("froze") the pre-trained weights up to the fourth convolutional block and allowed fine-tuning of the weights in all remaining layers (starting from the last convolutional block, named stage "conv5"). Finally, we used a fully-connected architecture again as the output submodel, with the same structure used for the data modality submodels. The next two sections provide more details on the data fusion and output layers.

### Multimodal data fusion layer

MultiSurv’s data fusion layer reduces the set of feature representation vectors to a single fusion vector, used as the input to the subsequent module. Let **Z** = [**z**_1_,..,**z***_n_*] be the matrix composed of the feature representation vectors, with **z*_l_*** ∈ ℝ*^m^* containing the feature representation of the *l^th^* input data modality. MultiSurv’s multimodal data fusion layer yields a compact representation vector **c** ∈ ℝ*^m^*, computed as the row-wise maximum of **Z**:

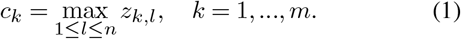

The fused feature representation thus corresponds to the maxima over the different data modalities. We also tested several alternative schemes, described in Suppl. Note 2, but found no improvement in performance.

### Discrete-time survival model formulation

MultiSurv is a fully parametric discrete-time survival model parameterized by a deep neural network. This formulation overcomes the proportionality constraint of CPH-based models and can be trained using stochastic gradient descent (SGD). Briefly, we assume that the follow-up time is discrete and let {*t*_1_, *t*_2_,…, *t_p_*} be the set of upper limits of *p* left-closed and right-open time intervals. For a given study subject, the hazard function *h_j_* defines the probability that the event of interest is observed in interval *j*, given that the subject has survived at least until the beginning of the interval. We used the negative of the log likelihood as the loss function to train the model. The log likelihood for time interval *j* is:

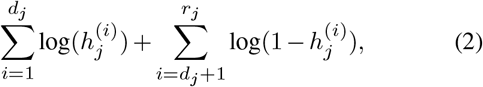

where 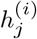 is the hazard probability for the *i*^th^ subject during time interval *j*. There are *r_j_* subjects at risk during interval *j* (with event or censoring time later than the beginning of the interval) and the first *d_j_* subjects experience the event during this interval. The total loss is the sum of the losses for each time interval [27]. Intuitively, the first term in Eq. (2) serves to encourage the model to increase, at each time interval *j*, the predicted hazard rate *h_j_* of the *d_j_* patients whose death occurred within that interval. The second term encourages the model to increase the predicted survival probability (1 − *h_j_*) for all *r_j_ − d_j_* patients who survived the interval. With this second term, in addition to uncensored patients, the loss leverages the information provided by censored patients, namely the fact that they are known to have survived time intervals earlier than their recorded censoring time.

MultiSurv uses a modification of the survival model implementation in [27], which was previously employed for simulated data and for life expectancy prediction of hospitalized patients from low-dimensional data. Briefly, MultiSurv’s prediction submodel (which takes as input the compact fusion vector generated by the fusion layer) is defined with an output layer containing *p* units, one for each time interval. A sigmoid activation function converts each unit’s output to a predicted conditional probability of surviving the respective time interval (corresponding to the complement of the conditional hazard rate in that interval, or 1 − *h_j_* for interval *j*). Patient *i’s* predicted probability of surviving through the end of time interval *j* is given by:

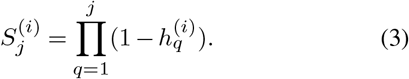

The loss function is a reformulation of Eq. (2) divided by study subject rather than by time interval, which facilitates implementation of the training procedure with mini-batches of patients.

Motivated by the idea that encouraging similarity between different feature representations may facilitate data fusion, we experimented with an auxiliary loss penalizing dissimilarity between the data modality representations. This auxiliary loss consisted of the average cosine distance between pairs of input data modality feature representations. The final loss was a weighted sum of main and auxiliary losses. Adding this auxiliary loss did not yield any improvement in performance, however, so we did not include it in the final MultiSurv configuration.

### Model training

Data from individual patients were randomly split into training (80%), validation (10%), and test (10%) datasets, stratified by cancer type. The validation set was used to assess the performance during iterative model development. We trained the models using Adam stochastic gradient descent optimization [43] in the PyTorch v1.4.0 implementation with default settings except for the learning rate. Initial learning rates were chosen using a learning rate range test in a pre-training run, performed by monitoring training loss over a linear range of learning rate values [44]. The resulting start values for the learning rate used for model training were between 0.001 and 0.005. These values were plugged into a scheduler set to reduce the learning rate upon learning stagnation: typically, a reduction by a factor of two after 10 epochs with no increase in validation performance. We also employed early stopping to save model states upon stagnation of learning (specifically, before registering an increase in validation loss). Models trained from scratch generally converged after less than 50 epochs of training.

### Model evaluation

We used two different time-dependent metrics to assess model performance. The first is the time-dependent concordance index, C^td^ [35], which is an extension of the widely used Harrell’s concordance index or C-index [37]. Coxian (CPH-based) methods are not designed to actually define the time-dependent baseline hazard term, but it can be estimated from the data to obtain additional information. By definition, Coxian methods have the same C-index at all predicted follow up times, since the time-dependent baseline hazard does not depend on the patient features. This means that the differences in predictions between individual patients are proportional and, hence, their predicted survival curves do not cross. Consequently, the survival probability rankings used to compute the C-index do not change over time. Having the same C-index at all predicted follow up times also means that C^td^ values for these methods are equal to their C-index values. The second metric we use is the integrated Brier score (IBS), which quantifies the average squared distances between observed survival status and predicted survival probabilities [36]. To calculate the IBS, we define a set of 100 equidistant time points between the minimum and maximum event times in the test dataset. We dropped the last quartile of time points, since the IBS typically becomes unstable at the latest time points.

All presented results were obtained using the test set. To avoid biased evaluation results, the test set remained unused and hidden until the final evaluations. In addition, we report 95% two-sided confidence intervals obtained using the non-parametric percentile bootstrap method with 1,000 samples from the test set. Since the data in the test dataset produces a 20-year long Kaplan-Meier estimate, for direct comparison we plot MultiSurv’s output up to 20 years post diagnosis (rather than the 30-year period predicted by MultiSurv).

### Previously published methods

We evaluated four previously published methods in order to establish baseline performance values that MultiSurv’s results could be directly compared to. The first is the classical CPH model [6]. The second is DeepSurv [13], a modern non-linear, DL based CPH method. Yet another baseline method is DeepHit [25], as a representative of the DL-based non-proportional methods. Finally, we used the random survival forest (RSF) method [30], a non-Coxian, non-proportional flexible alternative to the DL-based discrete-time approaches.

### Software and hardware

The software was developed using Python v3.6.8 (Anaconda v4.3.34 distribution). MultiSurv models were trained using PyTorch v1.4.0 [45] (with cuda v10.1.243 and cudnn v7.6.3) on a workstation equipped with an Nvidia GeForce GTX 1070 graphics processing unit (GPU) and a server equipped with two Nvidia GeForce RTX 2070 GPUs. The CPH and RSF baseline models were fit using the Python software packages lifelines v0.23.8 [41] and pysurvival v0.1.2 [46], respectively. The two DL-based baseline models, DeepSurv and DeepHit, were trained using the Python software package pycox v0.2.0 [23]. For the employed metrics, C^td^ and IBS, we used the implementations in pycox v0.2.0 [23]. To embed the representation vectors from the original 512-dimensional space into a two-dimensional space using the t-distributed Stochastic Neighbor Embedding algorithm (t-SNE) [38], we used the implementation in scikit-learn v0.22.1 [47], with a perplexity of 50 and otherwise default values. In addition, we relied on several other Python software packages to build important functionality, most notably NumPy v1.18.1 [48], SciPy v1.4.1 [49], and pandas v1.0.1 [50].

## Data Availability

All data used in this work are from The Cancer Genome Atlas (TCGA) program. The used datasets are available publicly from the National Cancer Institute (NCI) Genomic Data Commons (GDC) information system, accessible at the GDC Data Portal.

https://portal.gdc.cancer.gov/

## ACKNOWLEDGEMENTS

L.A.V.S. acknowledges the Chica and Heinz Schaller Foundation for funding through an individual fellowship. We thank Prof. Dr. med. Peter Sinn for help with histology slide images and the BMCV group members for helpful discussions. This preprint is formatted using a LATEX template by Ricardo Henriques.

